# Visible light optical coherence tomography of peripapillary retinal nerve fiber layer reflectivity in glaucoma

**DOI:** 10.1101/2021.08.31.21262930

**Authors:** Weiye Song, Sui Zhang, Yumi Mun Kim, Natalie Saldlak, Marissa G. Fiorello, Manishi Desai, Ji Yi

## Abstract

**Purpose:** To evaluate the clinical utility of visible light optical coherence tomography (VIS-OCT) and to test whether VIS-OCT reflectivity and spectroscopy of peripapillary retinal nerve fiber layer (pRNFL) are correlated with severity of glaucoma, compared with standard-of-care OCT thickness measurements.

**Design:** Cross-sectional study.

**Method:** Fifty-four eyes from three groups of subjects (normal, glaucoma suspect, and glaucoma) were scanned by a custom-designed dual-channel device that simultaneously acquired visible (VIS-OCT) and near-infrared OCT (NIR-OCT) images. A 5×5 mm^2^ scan was taken of the peripapillary nerve fiber layer (pRNFL).

The pRNFL reflectivity was calculated for both channels and the spectroscopic marker was quantified by pVN, defined by the ratio of VIS-OCT to NIR-OCT pRNFL reflectivity. The results were compared with ophthalmic exams and clinical Zeiss OCT measurements. Mixed linear model was used to evaluate the association of imaging markers with glaucoma severity.

**Results:** VIS-OCT pRNFL reflectivity significantly, se uentially declined from normal to suspect to glaucoma (3.42 ± 0.35, 2.58 ± 0.37, 2.16 ± 0.39, mean ± SD), as did NIR-OCT pRNFL reflectivity (2.46 ± 0.24, 2.27 ± 0.24, 1.96 ± 0.25). The pVN also had the same decreasing trend among three groups (1.39 ± 0.08, 1.14 ± 0.08, 1.08 ± 0.10). Normal and suspect eyes were significantly different in VIS-OCT pRNFL reflectivity (p=0.002), pVN (p<0.001) but not in NIR-OCT pRNFL reflectivity (p=0.14), circumpapillary RNFL (cpRNFL) thickness (p=0.17), or macular ganglion cell layer and inner plexiform layer (GCL+IPL) thickness (p=0.07) in the mixed linear model.

**Conclusion:** VIS-OCT pRNFL reflectivity and pVN was more sensitive in separating suspect eyes from normal ones than clinical OCT thickness measurements. VIS-OCT pRNFL reflectivity and pVN could be a useful metric in early detection of glaucoma upon further longitudinal validation.

## Introduction

Glaucoma is a leading cause of blindness worldwide, and the second leading cause of blindness in the United States^1,2^. Clinically, glaucomatous optic neuropathy is characterized by thinning in the peripapillary retinal nerve fiber layer (RNFL) and reduction in sensitivity in visual field testing^3–5^. Early detection of the structural damage to retinal ganglion cells (RGC) and their axons in the RNFL and within the optic nerve head is important for timely diagnosis, treatment and vision preservation. Optical coherence tomography (OCT)^6^ has been the standard of care imaging modality to quantitatively measure the structural change of RNFL. Peripapillary RNFL and macular RGC thinning are routinely used in diagnosing glaucoma^7–10^, but improvements in the sensitivity of these structural measures would be beneficial.

Laboratory studies have shown that changes in the RGC axon cytoskeleton can be detected by RNFL spectroscopic contrast and that these features may represent nanoscale change in microtubules prior to axon loss and consequent RNFL thinning^14–16^. Visible light OCT (VIS-OCT) is a recent development that uses shorter wavelengths than the near infrared (NIR) wavelength used in standard OCT devices^17–19^. Initial data show that the spectral contrast between VIS-OCT and NIR-OCT may reveal information from which nanoscale structural features can be quantitatively inferred^20–23^ at a scale beyond the image resolution by standard OCT images^24–25^ This approach, when applied to clinical reflectance spectroscopy, may provide better sensitivity on RNFL alterations in early glaucoma.

We recently developed a dual-channel clinical OCT device that can simultaneously acquire VIS-OCT and NIR-OCT using a robust fiber optics design^20^. A novel metric (*i.e*. pVN) was defined from reflectance spectroscopy, consisting of the intensity ratio between VIS-OCT and NIR-OCT images. We identified changes in RNFL of ocular hypertensive mice using pVN^26^. The goal of the present study was to compare our two-channel VIS-OCT metrics to standard OCT outcomes among normal, glaucoma suspect and glaucoma patients in a cross-sectional study.

## Methods

### Study design and subject

This cross-sectional study was conducted at Boston Medical Center eye clinics, from March 2019 to January 2020, whose Institutional Review Board reviewed and approved the study. The study was compliant with Health Insurance Portability and Accountability Act and adhered to the tenets of the Declaration of Helsinki. Written informed consent was obtained from all subjects before participation. Inclusion criteria were age ≥40 years, and best-corrected visual acuity better than 20/40. Exclusion criteria included best-corrected visual acuity worse than 20/40 in either eye, previous intraocular surgery other than uncomplicated cataract surgery, angle-closure glaucoma and other known retinal conditions. Cataracts were evaluated using the Lens Opacification System II (LOCS II) based upon color and opalescence^27^. The system uses a 4-point grading system with increasing number consistent with increasing maturity. Severe cataract graded more than 2+ were excluded.

Study subjects were recruited during their standard-of-care visit. Normal eyes were defined as having a normal appearance of the optic nerve and by stereoscopic optic disc photograph assessment at the point of care (subjective assessment of vertical cup-to-disc ratio, VCDR), as well as having no history of IOP above 22 mm Hg. Glaucoma suspect eyes were defined as having optic disc appearance judged potentially to represent glaucomatous optic neuropathy on stereoscopic optic nerve examination by M. Desai, but normal visual field (VF) test by Zeiss 24-2 threshold test: glaucoma hemifield test (GHT) either within normal range or borderline, and a pattern standard deviation index p < 5%. Glaucomatous eyes were defined as having optic disc appearance judged to be compatible with glaucoma and reliable, abnormal VFs: GHT result outside normal limits or a pattern standard deviation index < 0.05%.

All the subjects went through ophthalmic examination including tonometry, stereoscopic optic disc assessment, and clinical OCT imaging (Cirrus, Zeiss, Germany). RNFL thickness at peripapillary and macular region, average cup-disc ratio (CDR) and VCDR were quantitatively derived from standard OCT. Humphrey central 24-2 threshold tests were taken for suspect and glaucoma subjects and the mean deviation (MD) and pattern standard deviation (PSD) were recorded. After the clinical and ophthalmic examination, dual-channel VIS-OCT were performed subsequently, and RNFL reflectivity metrics were quantified. The patients were imaged by trained technicians.

### Research dual-channel VIS-OCT device

A custom-built dual-channel VIS-OCT device was used to acquire optic disc cubes from every subject (Fig. 1a). The visible channel used 545-580 nm light as the light source, and the NIR channel used wavelength at. Each scanning cube had 256 B-scan frames, and each B-scan had 512 A-lines. Each scan covered an approximately 5×5 mm^2^ area. Further technical details of the device were described in our previous publications^20^. The visible light channel used a supercontinuum source (SuperK Extreme, EXU-OCT-6, NKT, Denmark), and NIR channel used a superluminescent laser diode (SLD-CS-371-HP3-SM-840-I, Superlum, Russia). A custom-built spectrometer was used for VIS-OCT, and a commercial one (CobraS 800, Wasatch Photonics, US) for NIR-OCT. The laser powers on pupil were 0.25 mW and 0.9 mW for VIS- and NIR-OCT.

**Fig 1.**
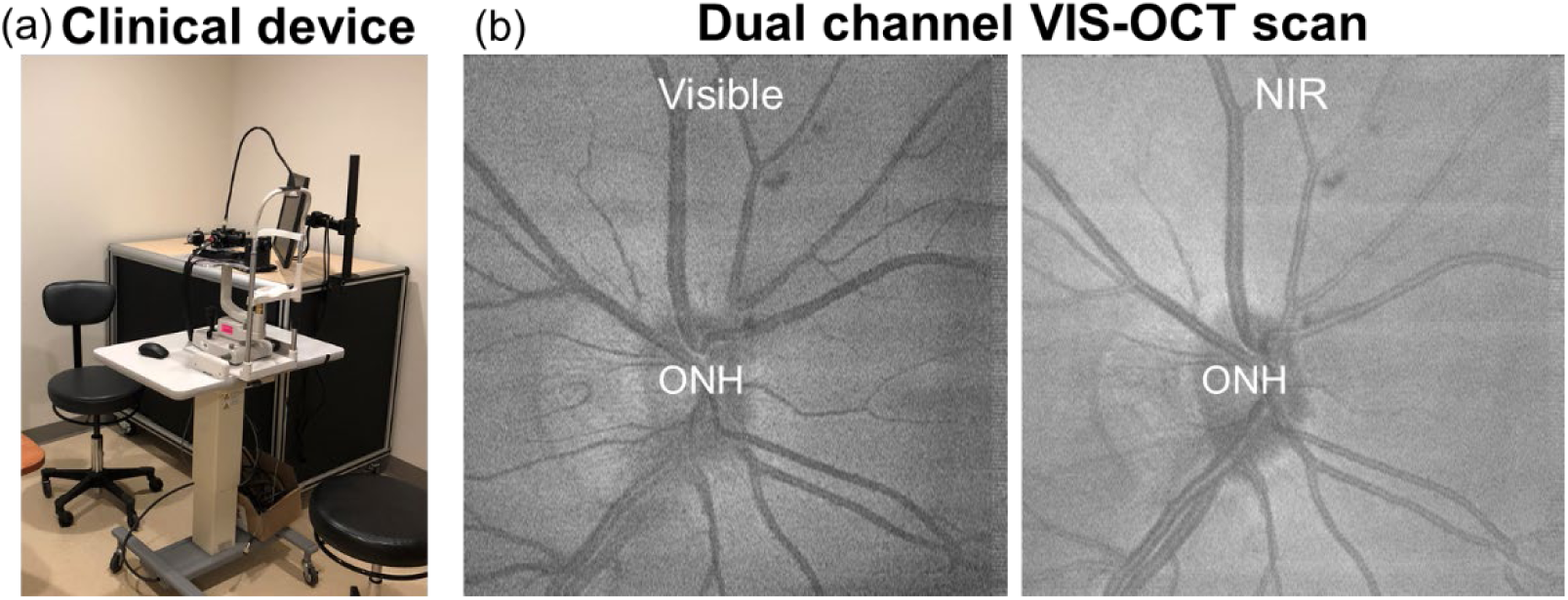
(a) Photograph of dual-channel VIS-OCT device. (b) *En face* projections of the optic disc scan in the visible and NIR channels, respectively.

We installed an electric tunable lens to accommodate the spherical refractive error. The fellow eye was used for fixation. The NIR channel was used for the initial alignment, then visible light channel was turned on for fine tuning and simultaneous dual-channel imaging. The acquisition time was 2.6s at 50 kHz A-line speed. Image examples are shown in Fig. 1b.

### RNFL reflectivity metrics and processing

All the image processing was based on the linear scale of OCT images. The pRNFL reflectivity metrics included VIS pRNFL-R (**p**eripapillary **RNFL r**eflectivity in VIS-OCT), NIR pRNFL-R (**p**eripapillary **RNFL r**eflectivity in NIR-OCT), pVN (ratio of **V**IS pRNFL-R and **N**IR pRNFL-R). The processing for each parameter is described below.

The manual segmentation was performed on NIR-OCT images to label the retinal surface, the bottom edge of RNFL and RPE. Dense points were first marked on the boundaries in ImageJ, and linear interpolations were performed within B-scans and subsequently along the slow direction. After the segmentation on NIR-OCT images, we registered VIS-OCT and NIR-OCT images and applied the same segmentation to both channels. RNFL and sub-RNFL layer were isolated as illustrated in Fig. 2. The noise background, *I*_*noise*_, was first obtained by averaging the signal within 30-40μm above the retina surface and subtracted from the image cube. In order to quantitate RNFL reflectivity, the following processing was performed. *En face* regions of interest (ROIs) were manually selected to avoid the major vessels. OCT signals within RNFL were summed, normalized by the thickness per A-line, and then averaged within areas in all ROIs.

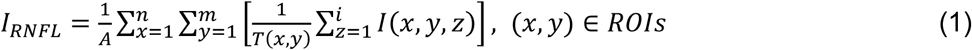

**Fig. 2.**
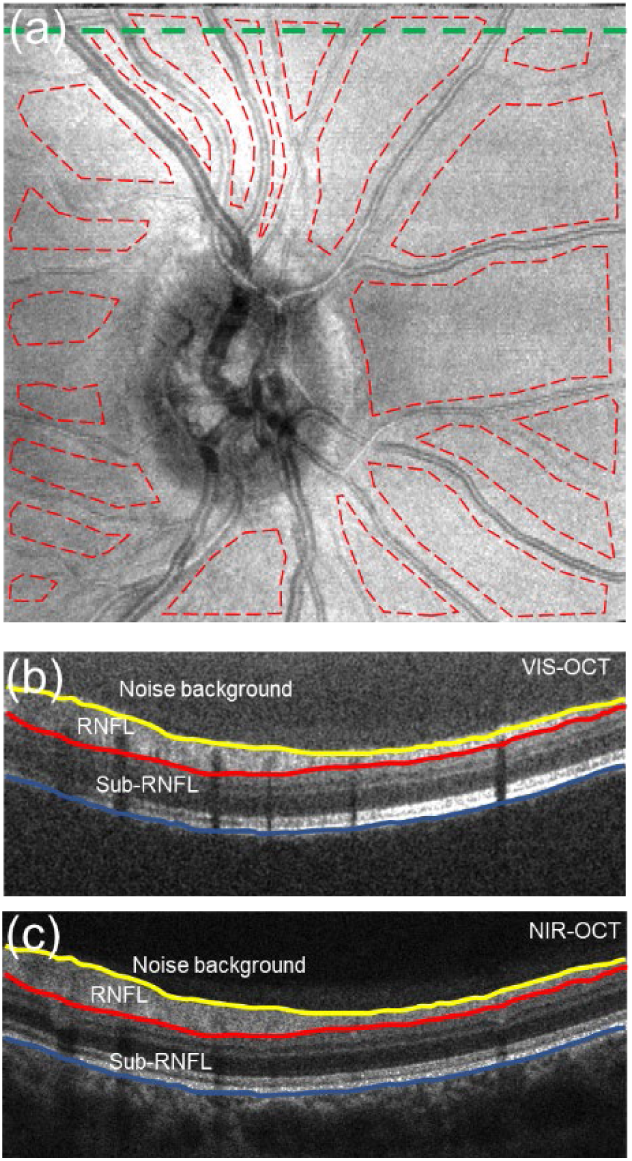
(a) Illustration of the *en face* ROIs selection. Red dashed line outlines the ROIs. (b-c) Segmentation for RNFL and sub-RNFL layer in VIS-OCT and NIR-OCT, respectively. B-scans in both channels were registered so that the same segmentation could be applied. B-scans were taken from the green dashed line in panel (a).

where *I(x,y,z)* is the cubic OCT signal, and *I*_*RNFL*_ is the RNFL signal. *A* is the total *en face* area of the selected ROIs, and *T*(x,y) is the RNFL thickness at each A-line. In order to account for the signal variation between eyes, we used the same method to calculate *I*_*sub-RNFL*_ within sub-RNFL layer and quantitate RNFL reflectivity by,

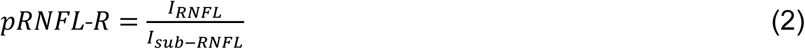

We then termed pRNFL reflectivity by VIS pRNFL-R and NIR pRNFL-R in the visible and NIR channels, respectively. The spectroscopic reflectivity contrast was further quantified by pVN defined below,

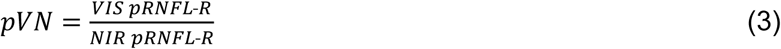

### Statistical analysis

Chi-square or one-way ANOVA were used to test the significance among three groups for categorical or continuous variables in the demographic information and ophthalmic exams. For variables by Zeiss OCT and dual-channel VIS-OCT, Spearman rank tests or two sample T-test were used to evaluate the association with three groups of subjects. Receiver operating characteristic (ROC) analysis was used to evaluate detection accuracy based on univariate logistic prediction. Mixed linear regression models were used to account for inter-subject variations considering some subjects had both eyes imaged. Negative logarithmic transformation was applied to mean deviation (MD) to improve the normalcy distribution in the mixed linear regression. When MD>0, MD was set to be the maximum negative MD value in this study. Adjusted average and 95% confidence interval (CI) were calculated by mixed linear model to evaluate the impact of severity and cataract to individual variables. All statistical analysis was performed using SAS 9.4.

## Results

### Characteristics of study subjects

The study included 36 subjects (11 normal, 12 suspect, 13 glaucoma) and 54 eyes (20 normal, 17 suspect, 17 glaucoma) were successfully imaged with completed datasets. Examples of pRNFL reflectivity, Zeiss OCT, and 24-2 VF tests were shown in Fig. 3. There is no statistical difference among groups on age, gender, race and ethnicity distributions (Table 1). By examining the demographic and ocular characteristics of study eyes (Table 2), there was moderate increasing percentage of cataract from normal, suspect to glaucomatous eyes. Zeiss OCT parameters were monotonically related among three groups, as well as VIS pRNFL-R, NIR pRNFL-R, and pVN.

**Fig. 3.**
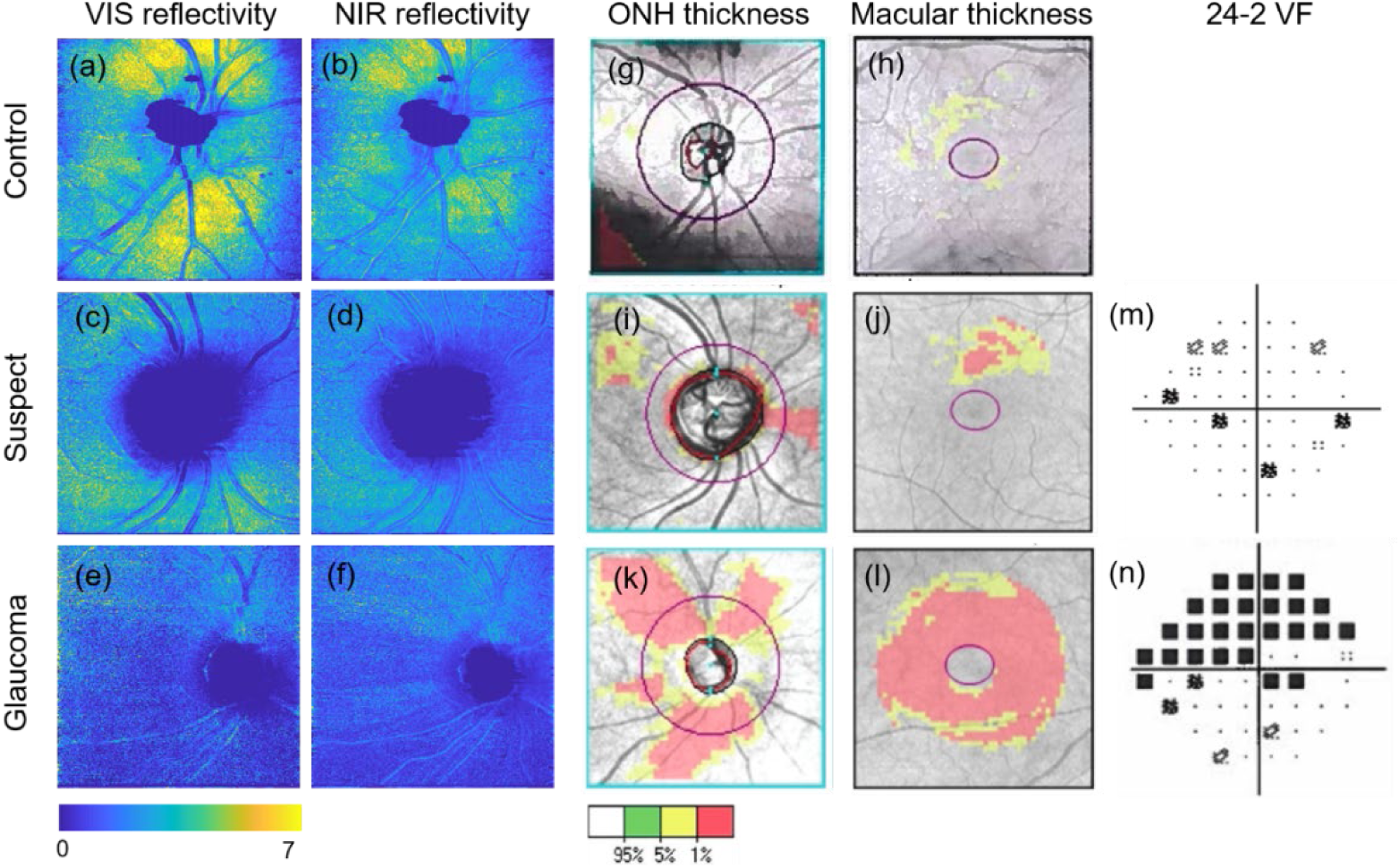
Comparison of pRNFL reflectivity in visible and NIR channel, ONH and macular OCT scans, and visual field test in three examples, one from each group. (a-f) *En face* mapping of VIS-pRNFL-R and NIR-pRNFL-R from dual-channel OCT measurements. (g-l) ONH and macular cube scan by Zeiss Cirrus OCT with the thickness abnormality. (m-n) 24-2 pattern deviation map.

**Table 1.**
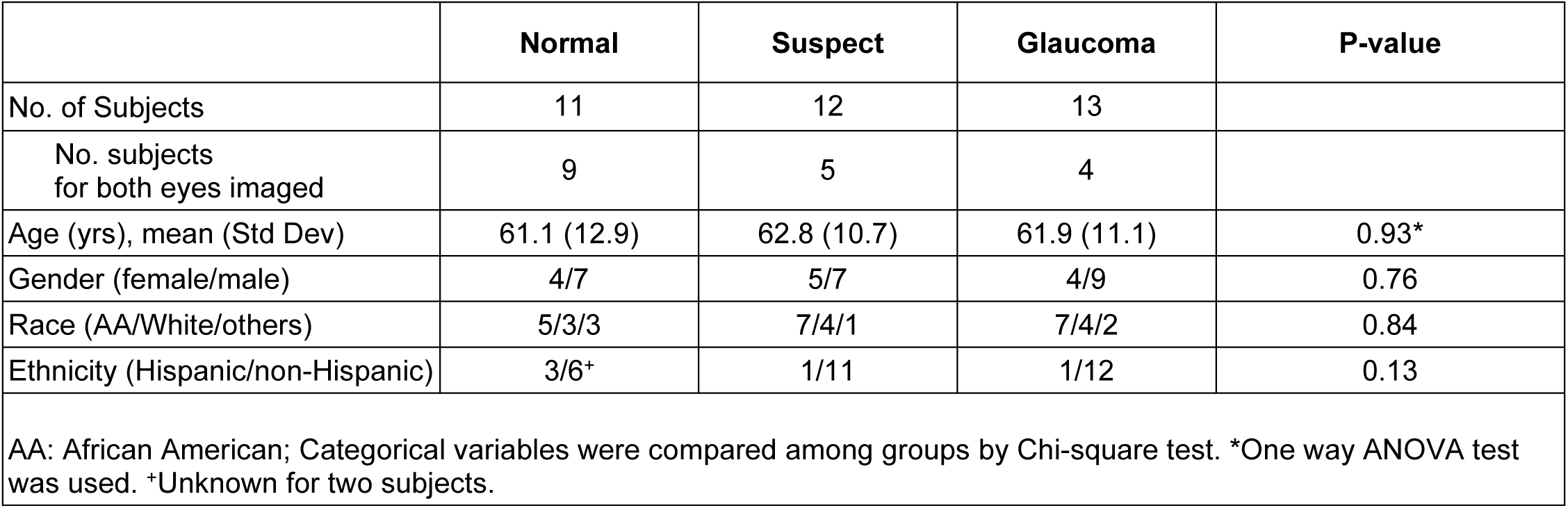
Demographic information of Study Subjects.

**Table 2.** Demographic and Ocular Characteristics of Study Eyes.

|  | Normal | Suspect | Glaucoma | P-value |
| --- | --- | --- | --- | --- |
| No. of eyes | 20 | 17 | 17 |  |
| Age (yrs), mean (Std Dev) | 60.2 (12.7) | 62.1 (9.8) | 61.9 (10.6) | 0.86<sup>+</sup> |
| Side (OS/OD) | 10/10 | 9/8 | 9/8 | 0.98<sup>^</sup> |
| Gender ratio (female/male) | 7/13 | 9/8 | 5/12 | 0.34<sup>^</sup> |
| Ophthalmic Exam |  |  |  |  |
| Cataract (Yes/No) | 11/9 | 11/6 | 15/2 | 0.09<sup>^</sup> |
| Pseudophakic lens (Yes/No) | 5/15 | 4/13 | 2/15 | 0.33<sup>^</sup> |
| IOP (mmHg), mean (Std Dev) | 15.3 (2.0) | 15.4 (3.7) | 14.1 (3.7) | 0.52<sup>+</sup> |
| Cylinder, mean(Std Dev) | 1.0 (0.7) | 0.4 (0.4) | 1.0 (0.5) | 0.75<sup>+</sup> |
| Visual field test |  |  |  |  |
| 24-2 MD (dB), mean (Std Dev) | - | -0.6 (1.2) | -7.0 (5.4) | **0.0002\*** |
| 24-2 PSD (dB), mean (Std Dev) | - | 1.8 (0.4) | 7.1 (5.0) | **0.0004\*** |
| Zeiss OCT |  |  |  |  |
| GCL+IPL (um), mean(Std Dev) | 77.8 (8.5) | 72.6 (8.6) | 64.4 (11.3) | **0.0006#** |
| cpRNFL (um), mean(Std Dev) | 89.8 (9.4) | 84.4 (14.3) | 67.9 (14.0) | **<0.0001#** |
| Averaged CDR, mean(Std Dev) | 0.5 (0.2) | 0.7 (0.1) | 0.7 (0.1) | **<0.0001#** |
| VCDR, mean (Std Dev) | 0.5 (0.2) | 0.7 (0.1) | 0.7 (0.1) | **<0.0001#** |
| Dual-channel VIS-OCT |  |  |  |  |
| VIS pRNFL-R, mean(Std Dev) | 3.4 (0.9) | 2.5 (0.7) | 2.0 (0.6) | **<0.0001#** |
| NIR pRNFL-R, mean(Std Dev) | 2.5 (0.5) | 2.2 (0.5) | 1.9 (0.4) | **0.0003#** |
| pVN, mean(Std Dev) | 1.4 (0.2) | 1.1 (0.2) | 1.0 (0.1) | **<0.0001#** |
| IOP: intraocular pressure; MD: mean deviation; PSD: pattern standard deviation; GCL+IPL: ganglion cell layer and inner plexiform layer thickness in macular scan; cpRNFL: circumpapillary retinal nerve fiber layer thickness. CDR: cup-disc ratio; VCDR: vertical cup-disc ratio. Dual-channel VIS-OCT markers are averaged values from all ROIs per eye. <sup>+</sup>One way ANOVA was used. <sup>^</sup>Chi-square test was used for categorical variables. <sup>*</sup>Two sample T-test was used. <sup>#</sup>Spearman rank tests were used to evaluate the changes of variables among three groups. |  |  |  |  |

### Clinical performance for two channel VIS-OCT markers

Both VIS pRNFL-R and NIR pRNFL-R decreased from normal to suspect to glaucoma groups (Fig. 4a, 4b). The spectroscopic marker pVN also decreased, suggesting that the change of RNFL reflectivity is more dominant in visible channel than NIR (Fig. 4c). Between suspect and glaucomatous eyes, both VIS pRNFL-R and NIR pRNFL-R were significant.

**Fig. 4.**
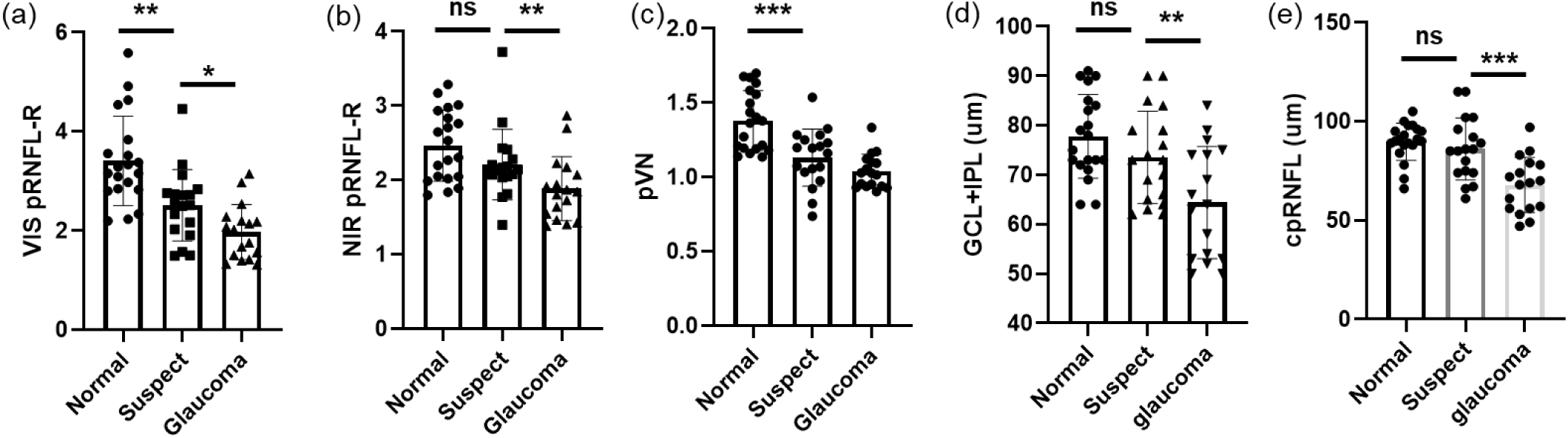
(a-c) Peripapillary RNFL reflectivity in VIS- and NIR-OCT channels, and pVN ratio in three groups of subjects. (d-e) Zeiss OCT thickness in three groups of subjects. Two sample T-test was used to test significance within two groups. ^*^p<0.05, ^**^p<0.01, ^***^p<0.001

In comparison with OCT thickness measurement, pVN and VIS pRNFL-R had higher significance in separating normal and suspect eyes. Neither GCL+IPL nor cpRNFL was significantly different between normal and suspect eyes (Fig. 4d-4e). ROC analysis confirmed this observation that pVN and VIS pRNFL-R had the higher AUC in comparison with GCL+IPL in separating suspect from normal eyes (Table 3). At the same time, VIS-OCT markers had lower AUC values than cpRNFL in differentiating suspect and glaucoma eyes.

**Table 3.** Area Under the Curve by ROC analysis.

|  | Normal vs. Suspect | Suspect vs. Glaucoma | Normal vs. Suspect + Glaucoma |
| --- | --- | --- | --- |
| VIS pRNFL-R, (95% CI) | 0.824(0.684-0.963) | 0.723(0.547-0.900) | 0.882(0.793-0.972) |
| NIR pRNFL-R, (95% CI) | 0.638(0.449-0.827) | 0.723(0.540-0.907) | 0.731(0.590-0.872) |
| pVN, (95% CI) | 0.788(0.642-0.935) | 0.671(0.475-0.867) | 0.872(0.781-0.963) |
| GCL+IPL, (95% CI) | 0.663(0.484-0.842) | 0.690(0.507-0.873) | 0.730(0.596-0.864) |
| cpRNFL, (95% CI) | 0.650(0.455-0.845) | 0.801(0.651-0.951) | 0.773(0.642-0.903) |
| Receiver operating characteristic (ROC) analysis is based on univariate logistic prediction. |  |  |  |

### Correlation between dual-channel VIS-OCT and standard Zeiss OCT

Spearman correlation tests showed that all markers from dual-channel VIS-OCT had excellent correlation with cpRNFL (Table 4). VIS pRNFL-R and pVN were highly correlated with GCL+IPL, as well as MD. Above descriptive population statistics suggested that VIS-pRNFL-R and pVN had better association with VF and Zeiss OCT measurements. To further test VIS-pRNFL-R and pVN’s correlation with glaucoma severity, a mixed linear regression model was used to account for individual variation because some of subjects have both eyes imaged. In the model where pVN was the dependent variable, cpRNFL and MD were independent variables, both cpRNFL (p=0.003) and MD (p=0.033) were significant. If we consider only within glaucoma eyes, both cpRNFL (p=0.000) and MD (p=0.006) remained significant when VIS pRNFL-R was dependent variable. VIS pRNFL-R and pVN were consistently correlated with cpRNFL in different tests.

**Table 4.**
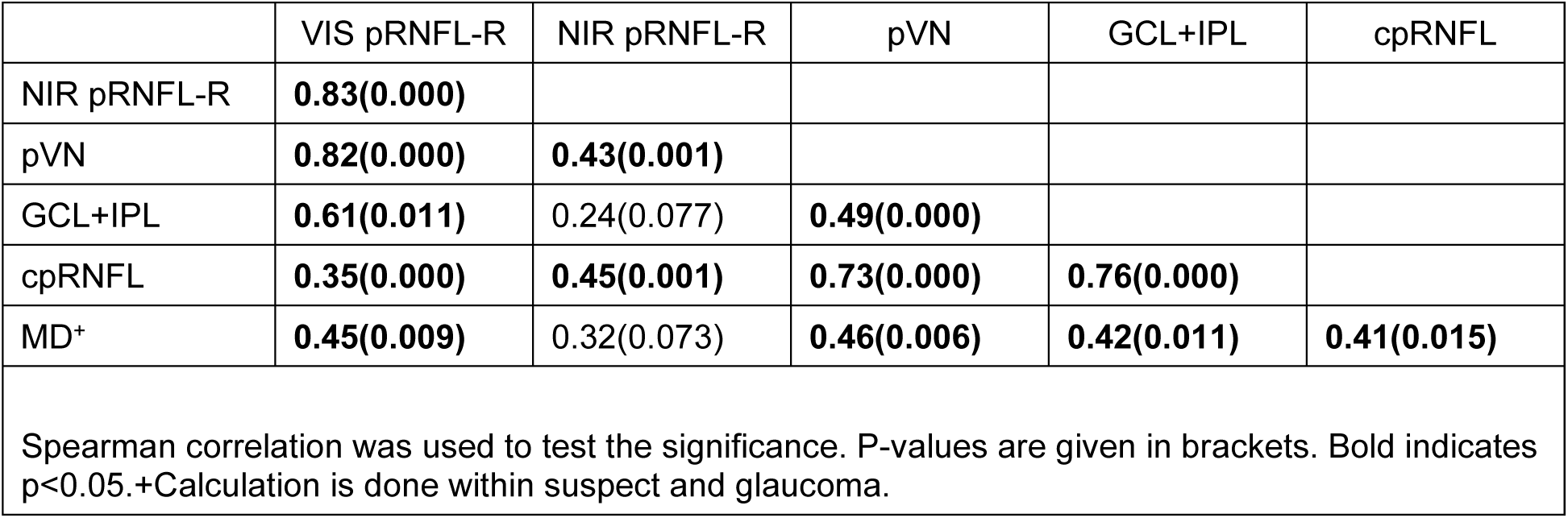
Correlation among dual-channel VIS-OCT, Zeiss OCT and visual field measurements.

### Cataract impact to imaging markers

We found that suspect and glaucomatous group had more eyes with cataract than the ones in normal group (Table 2), which could confound imaging markers. Mixed linear model regression was used to take glaucoma severity and presence of cataract as independent variables, where different imaging markers were dependent variables (Table 5). VIS pRNFL-R (p=0.002) and pVN (p<0.001) remained significant in separating normal and suspect eyes, and VIS pRNFL-R (p=0.043) and NIR pRNFL-R (p=0.017) were still significant in separating suspect and glaucoma eyes, with the presence of cataract. The pVN had higher significance in separating normal and suspect eyes than VIS pRNFL-R. Neither GCL+IPL nor cpRNFL significantly differentiated between normal and suspects eyes. Figure 5 further illustrates the adjusted averages (95% CI) for eyes in three groups with and without cataract. P-interaction suggested that cataract had no significant impact on the association between dual-channel VIS-OCT markers, Zeiss OCT thickness and glaucoma severity. The decreasing trends for all dual-channel VIS-OCT markers with increasing severity were consistent with and without cataract.

**Table 5.**
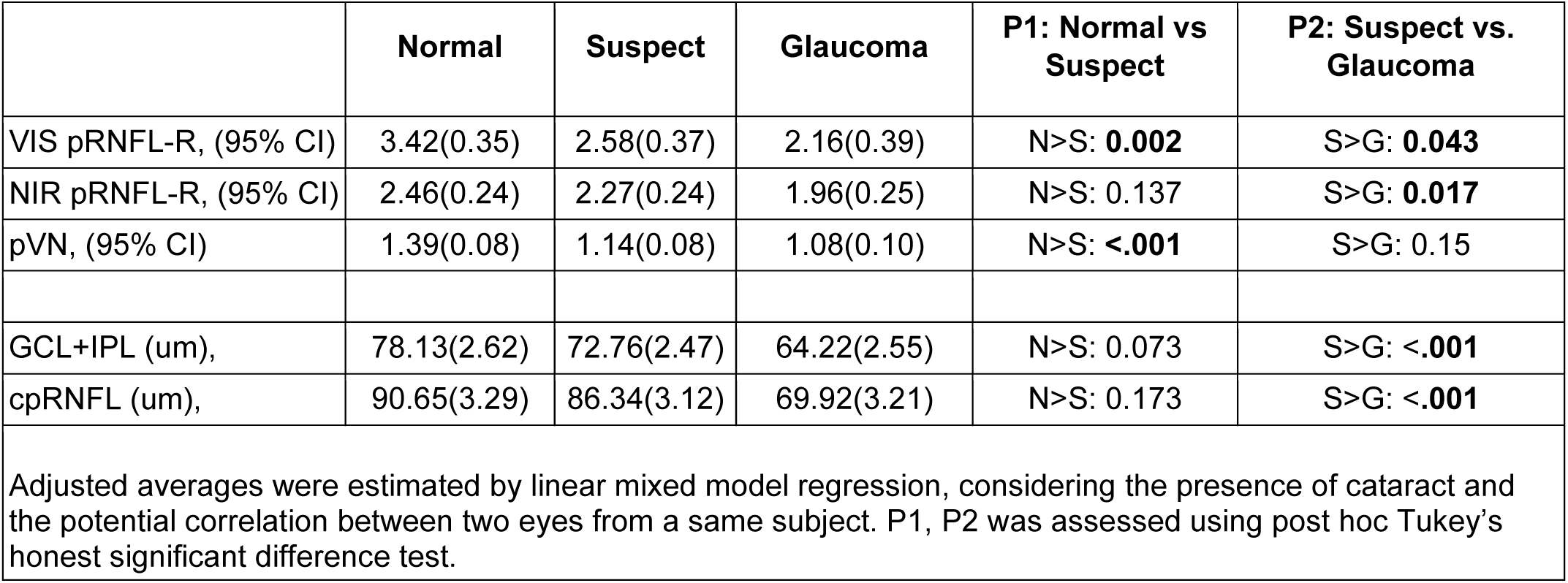
Adjusted average values by mixed linear model in three groups.

**Fig. 5.**
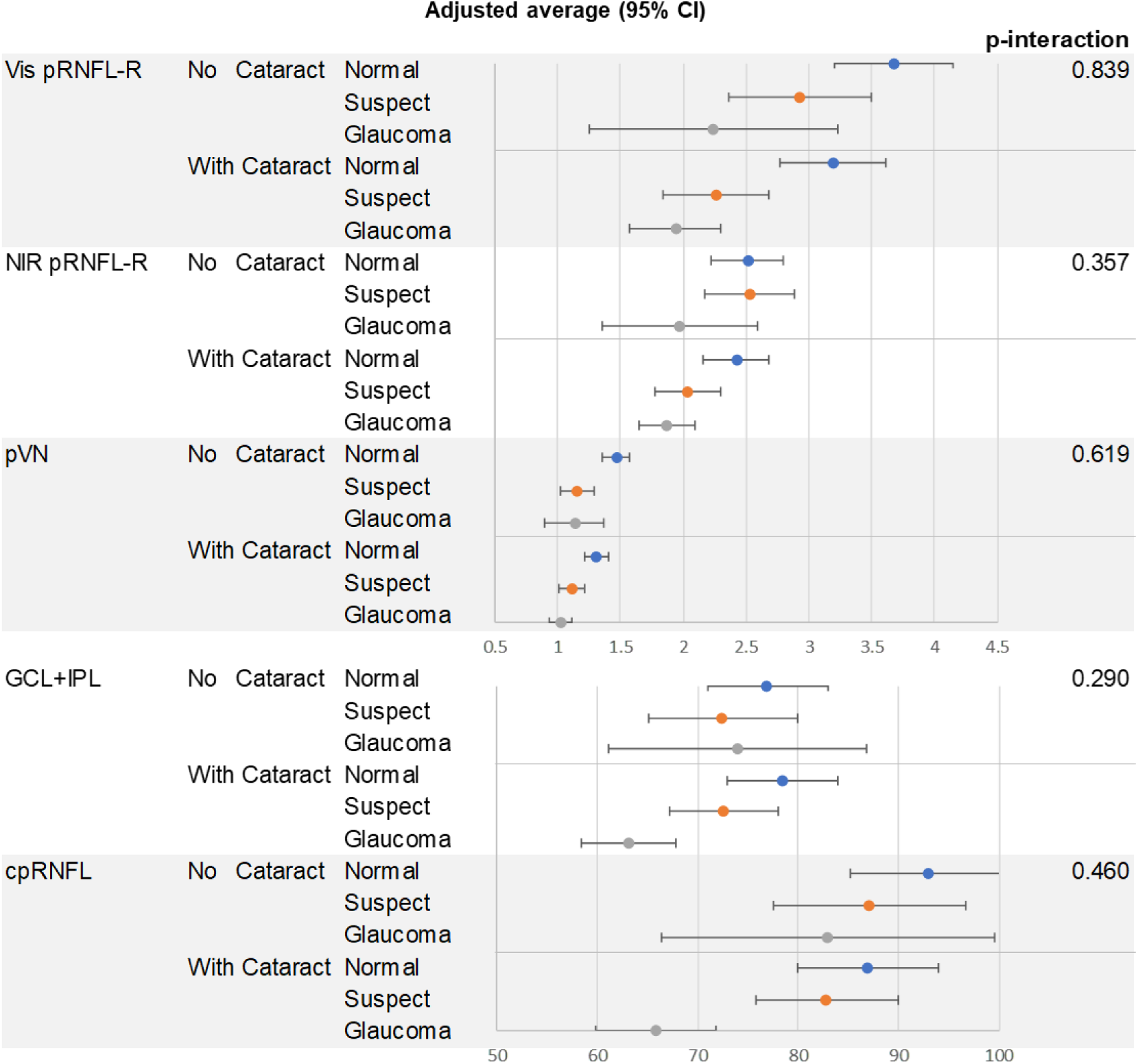
Adjusted average by mixed linear model regression, considering the potential correlation between two eyes from a same subject.

## Discussion

Our pilot study demonstrated the feasibility of VIS-OCT in peripapillary imaging in clinical subjects. The dual-channel device provided a simultaneous imaging on VIS-OCT and NIR-OCT, as well as reflectance spectroscopy by contrasting two channels. The dual-channel design allows ease of alignment using NIR light without excessive exposure of visible light before acquisition. We found a progressive decrease of both pRNFL reflectivity in VIS-OCT and NIR-OCT with increasing severity of glaucoma, and the same trend for pVN, indicating that reflectivity decreases more in the visible channel. Importantly, the VIS pRNFL reflectivity and pVN are more sensitive in separating glaucoma suspect from normal eyes than pRNFL and GCL+IPL thinning. We also showed that pVN has better significance than visible pRNFL reflectivity in differentiating suspect and normal eyes (Table 5), presumably due to the improved robustness from normalizing VIS-OCT by NIR-OCT.

The biophysical origin for the RNFL reflectance and spectroscopy has been extensively studied by Knighton and Huang *et al*. through a multispectral imaging microreflectometer^28^ Cytoskeletons in the axon of RGCs attribute to the reflectance signal, particularly microtubules^14^. Using an ocular hypertension glaucoma rat model, it was found that all cytoskeleton components were distorted and damaged, leading to the reduction of the RNFL reflectance signal^15^. This reduction preceded RNFL thinning^16^. A light scattering model elucidated that the loss of microtubules contributed to the spectroscopic change in RNFL, causing nonuniform changes in RNFL reflectance across wavelengths^15^. The RNFL reflectance was reduced in glaucomatous retinas more in visible than in NIR wavelengths. This is consistent from our data that pRNFL reflectivity in both channels exhibited declining trends with glaucoma severity, as did pVN. The reduction of pVN is also consistent with our *in vivo* study with an ocular hypertension mouse model using dual channel VIS-OCT^26^. It is important to note that the spectroscopic change is sensitive to loss of microtubules in the length scale of tens of nanometer, beyond the resolution capability of any *in vivo* ophthalmic imaging modalities. The superb sensitivity of using spectroscopy is indeed well documented in the biophotonics field^24,25,29,30^, supporting that RNFL reflectance spectroscopy could be an early detection method for glaucoma.

The existing clinical studies using RNFL reflectance in glaucoma are in general in line with our findings. A cross-sectional study showed that the reduction of RNFL reflectivity is associated with glaucoma, and could better distinguish pre-perimetric glaucoma from normal eyes^31^. However, the study used the summed intensity within RNFL and could be confounded by the RNFL thickness. Another longitudinal study reported a reduction in the RNFL reflectance may predict functional deterioration of glaucoma progression when combined with the rate of RNFL thinning^32^. Importantly, the data processing methods in these two studies were based on images taken from standard OCT devices. They likely used conventional OCT image intensity on a logarithmic scale, which could blunt the contrast and limit the sensitivity. In contrast, we used the linear scale images for calculating the reflectance signal. We performed a normalization to cpRNFL thickness and the sub-RNFL tissue intensity, and further calculated the pVN based on the ratio between two channels. These processing steps may contribute to better sensitivity, by removing the confounder of RNFL thinning and varying image quality.

Although the visible RNFL reflectivity and pVN have the best separations between normal and suspect eyes than than RNFL thinning. However, they may reach a floor effect ^33,34^ more quickly (*i.e*. less significant difference between suspect and glaucoma eyes) than RNFL thickness. This behavior implies that both metrics are earlier events than thinning, presumably due to the nanoscale sensitivity.

There are several limitations in our study. First, patient number in this pilot study is limited. Further validation will benefit from a larger number of subjects. Second, this is a cross-sectional study with no indication of whether VIS-OCT pRNFL reflectance and pVN can be predictive of progression. Third, patients with severe cataract were excluded, because of the unquantifiable VIS-OCT image quality. Due to the laser power limitation and shorter wavelength, VIS-OCT is more susceptible to scattering attenuation from cataract. Fourth, the current layer segmentation and the selection of ROIs were performed manually in this study for accuracy. Automatic software will be developed to rapidly perform the processing.

Future work includes continuing to improve the VIS-OCT device by having better light source stability, better detection sensitivity, optimizing/standardizing the imaging protocol, and streamlining the data processing.

Beyond the scattering spectroscopy exploited here, VIS-OCT enables high resolution^35,36^ and accurate microvascular retinal oximetry^37–40^. By using the technology offered by VIS-OCT, we can continue to enhance our ability to understand the sequence of physiologic changes in glaucoma in addition to improving our ability to detect abnormalities earlier. This technology also may have application in other retinal pathologies.

## Data Availability

The datasets generated during and/or analysed during the current study are available from the corresponding author on reasonable request.

## Acknowledgement

This study was supported by BrightFocus foundation (G2017077), NEI/NINDS R01NS108464. We acknowledge Dr. Harry Quigley, Pradeep Ramulu, and Thomas V. Johnston’s valuable discussion and comments for this manuscript.

